# Exploring the role of attention towards balance in chronic dizziness: Development of the Balance Vigilance Questionnaire (Balance-VQ)

**DOI:** 10.1101/2023.07.17.23292759

**Authors:** Toby J Ellmers, Elmar C Kal

**Author notes:** **Corresponding author:** Dr Toby J Ellmers.

## Abstract

**Background and Objectives:** Vigilance towards balance has been proposed to underpin various chronic dizziness disorders, including Persistent Postural Perceptual Dizziness (PPPD). The objective of this study is to develop (through patient input) a validated balance-specific measure of vigilance that comprehensively assesses the varied ways in which this construct may manifest.

**Methods:** We developed the Balance Vigilance Questionnaire (Balance-VQ) through patient and clinician feedback, designed to assess vigilance towards balance. We then validated the questionnaire in 497 participants consisting of patients diagnosed with chronic dizziness disorders (including 97 individuals diagnosed with PPPD) and healthy controls.

**Results:** The final 6-item Balance-VQ was shown to be a valid and reliable way to assess vigilance towards balance. Scores were significantly higher in individuals diagnosed with PPPD compared to controls. Although scores were also higher in the PPPD group compared to individuals with diagnosed vestibular disorders other than PPPD, Balance-VQ scores did not discriminate between the two groups when confounding factors were controlled for.

**Conclusions:** Our findings confirm that the Balance-VQ is a valid and reliable instrument for assessing vigilance towards balance. As symptom vigilance has been identified as a key risk factor for developing chronic dizziness following an acute neuro-otological insult, we recommend using the Balance-VQ as a screening tool in people presenting with such symptoms.

**Key Messages:**

- Vigilance towards balance has been proposed to underpin the development and maintenance of chronic dizziness disorders, such as Persistent Postural Perceptual Dizziness (PPPD).
- Clinically assessing balance vigilance is difficult, as no validated assessment method exists.
- Through feedback from patients and clinicians, we developed a new scale capable of assessing this construct: The Balance Vigilance Questionnaire (Balance-VQ).
- Our findings confirm that the Balance-VQ is a valid and reliable instrument for assessing vigilance towards balance.
- We recommend using the Balance-VQ as a screening tool in people at risk of developing, or currently presenting with, chronic dizziness.

## Introduction

Vigilant monitoring of body signals has been associated with the development and maintenance of a range of clinical disorders, including persistent postural-perceptual dizziness (PPPD).^1–5^ PPPD is a recently defined disorder of chronic dizziness, characterised by non-spinning vertigo, perceived/subjective unsteadiness and hyper-sensitivity to motion.^1^ Symptoms associated with PPPD often develop following acute (neuro-)vestibular insult.^1–3^ Whilst some degree of vigilance towards balance likely reflects an adaptive response to vestibular dysfunction and imbalance, a recent systematic review concluded that excessive monitoring of balance appears a strong risk factor for developing persistent dizziness symptoms.^4^

Although the specific mechanisms through which balance vigilance may contribute to dizziness remain unknown, some researchers have hypothesised that heightened monitoring of balance in PPPD leads to greater awareness of minor (otherwise subconscious) discrepancies between anticipated and actual postural feedback signals.^2, 3^ This mismatch may then lead to a distorted sense of imbalance and feelings of dizziness. This idea is supported by our recent experimental work highlighting how balance vigilance contributes to the formation of distorted perceptions of unsteadiness in healthy (older) adults without vestibular deficits.^6^ Balance vigilance has also been proposed to play a role in the maintenance of ‘unexplained dizziness’ in older adults.^6^ Here, dizziness is characterised by vague – and distorted – feelings of unsteadiness and imbalance, despite a lack of readily-identifiable neuro-otological dysfunction.^7, 8^

Comprehensive understanding of the specific role that balance vigilance plays in the maintenance of clinical balance disorders is therefore important for developing any future therapeutic strategies. However, to do so requires a uniformly used validated tool that specifically assesses balance vigilance. While a Body Vigilance Scale^9^ exists, this tool was developed to assesses generalised vigilance in panic disorder, and the amount of attention directed towards monitoring a broad range of sensations not inherently related to balance^a^ (e.g., upset stomach, heart palpitations, shortness of breath). As these broad items fail to distinguish between healthy controls and individuals with balance disorders, such as PPPD,^10^ there is a need to develop (through patient input) a validated balance-specific measure of vigilance that provides a comprehensive yet efficient assessment of the varied ways in which balance vigilance may manifest. Doing so will also help identify those most likely to benefit from specific, tailored therapeutic strategies. This is the aim of the present work.

## Methods

### Participants

555 participants (including healthy controls and individuals experiencing chronic and acute dizziness) were recruited from social support groups, community groups and patient support networks within the UK, North America and Australia. Due to the incidence peak of chronic dizziness in middle-and older-age,^11^ we excluded individuals aged <30 years of age. Participants were also excluded if they had been diagnosed with dementia or any other degenerative neurological disease. The final sample consisted of 97 individuals diagnosed with PPPD, 97 with a diagnosed vestibular disorder other than PPPD, and 303 controls without diagnosed neuro-otological dysfunction.^b^ Please see Table 1 for full demographic breakdown of the sample. Ethical approval was obtained from the local ethics committee, and the research was carried out in accordance with the Declaration of Helsinki. All participants provided written informed consent.

**Table 1.** Participant characteristics.

|  | **Vestibular**<br>**(N = 97)** | **PPPD**<br>**(N = 97)** | **Controls**<br>**(no neuro-otological dysfunction; N = 303)** |  |
| --- | --- | --- | --- | --- |
|  |  |  | *Control-dizzy (N=98)* | *Control-not dizzy (N=203)\** |
| **Background / General Health** |  |  |  |  |
| Age in years (mean ± SD [range]) | 62.8 ± 13.6<br>[31-87] | 47.2 ± 11.2<br>[30-76]<sup>b</sup> | 70.9 ± 10.3<br>[34-90]<sup>a</sup> | 68.2 ± 15.5<br>[30-90] |
| Female gender (n; %) | 86 (91%)<sup>b</sup> | 79 (82%)<sup>a</sup> | 80 (83%)<sup>a</sup> | 144 (71%) |
| Education Level – college / sixth form or above (n, %) | 87 (90%)<sup>a</sup> | 79 (82%)<sup>a</sup> | 81 (85%)<sup>b</sup> | 179 (89%)<sup>b</sup> |
| General health – self-reporting very good to excellent health (n; %) | 31 (32%)<sup>a</sup> | 19 (20%) | 52 (53%) | 126 (63%)<sup>b</sup> |
| Medication use >4 (n; %) | 14 (15%)<sup>a</sup> | 8 (8%) | 16 (16%) | 27 (13%) |
| Peripheral neuropathy (n; %) | 3 (3%)<sup>a</sup> | 2 (2%) | 10 (10%)<sup>a</sup> | 6 (3%)<sup>2</sup> |
| Diabetes (n; %) | 5 (5%)<sup>c</sup> | 6 (6%)<sup>a</sup> | 9 (9%) | 20 (10%)<sup>c</sup> |
| **Physical functioning** |  |  |  |  |
| Falls in past 12 months (n; %) | 41 (43%)<sup>a</sup> | 31 (32%)<sup>a</sup> | 43 (44%)<sup>a</sup> | 53 (26%)<sup>b</sup> |
| Balance problems (n; %) | 74 (79%)<sup>c</sup> | 84 (87%) | 50 (51%) | 58 (29%)<sup>e</sup> |
| Walking aid (n; %) | 12 (13%)<sup>c</sup> | 11 (11%) | 14 (14%)<sup>a</sup> | 14 (7%)<sup>b</sup> |
| ADL-assistance (n; %) | 4 (4%)<sup>b</sup> | 8 (8%) | 1 (1%)<sup>a</sup> | 0 (0%)<sup>d</sup> |
| **Psychological functioning** |  |  |  |  |
| Falls Efficacy Scale - International (7-28; mean ± SD [range]) | 12.0 ± 4.2<br>[7-28] | 13.1 ± 4.5<br>[7-23] | 11.1 ± 4.1<br>[7-24] | 9.3 ± 2.9<br>[7-27] |
| HADS-Anx (0-21; mean ± SD [range]) | 7.2 ± 4.4<br>[0-19] | 10.0 ± 5.4<br>[0-21] | 5.6 ± 3.7<br>[0-15] | 3.9 ± 3.3<br>[0-18] |
| Depression Diagnosis (n; %) | 44 (46%)<sup>b</sup> | 35 (36%) | 21 (21%) | 24 (12%)<sup>c</sup> |
| **Dizziness characteristics** |  |  |  |  |
| VSS – Total score (0-60; mean ± SD [range]) | 13.2 ± 10.3<sup>e</sup><br>[0-50] | 21.7 ± 10.9<sup>b</sup><br>[4-51] | 7.0 ± 6.4<sup>f</sup><br>[0-50] | 2.9 ± 3.1<sup>g</sup><br>[0-17] |
| VSS – Vertigo Subscale (0-32) (Mean ± SD [range]) | 8.2 ± 7.3<sup>e</sup><br>[0-31] | 13.6 ± 7.3<sup>b</sup><br>[1-31] | 2.9 ± 3.5<sup>f</sup><br>[0-28] | 0.9 ± 1.8<sup>f</sup><br>[0-15] |
| VSS – Arousal Subscale (0-28) (Mean ± SD [range]) | 5.0 ± 4.1<sup>e</sup><br>[0-20] | 8.1 ± 5.2<sup>b</sup><br>[0-23] | 4.0 ± 3.6<sup>f</sup><br>[0-22] | 2.0 ± 2.1<sup>f</sup><br>[0-8] |

**NB:** \* Missing data for two participants throughout, as they did not answer question on dizziness experience; <sup>a</sup>1 missing value; <sup>b</sup>2 missing values; <sup>c</sup>3 missing values; <sup>d</sup>4 missing values; <sup>e</sup>5 missing values; <sup>f</sup>10 missing values; <sup>g</sup>26 missing values;
**Abbreviations:** ADL: Activities of Daily Living; HADS-Anx: Anxiety subscale of the Hospital Anxiety Depression Scale; SD: Standard Deviation; VSS: Vertigo Symptom Scale;

### Development of the Balance Vigilance Questionnaire (Balance-VQ)

We adapted the existing Pain Vigilance and Awareness Questionnaire (PVAQ)^12^ to balance/dizziness through Patient and Public Involvement (PPI), conducted with fifteen individuals living with a variety of balance disorders (including PPPD, BPPV, undiagnosed chronic dizziness and older adults with generalised imbalance) and four experienced clinicians with expertise in treating balance disorders. This reiterative process involved first adapting individual items from pain to balance/dizziness, and then removing any items which were now deemed irrelevant. Discussions revealed a number of important constructs relevant to balance vigilance that were not captured in the original PVAQ. Items to capture these constructs were developed through discussions with the PPI members. Wording of each item was refined through further discussion, and feedback on each iteration of the Balance-VQ continued until consensus and agreement was reached.

This resulted in an 11-item Balance-VQ that was used for validation. This initial version of the scale can be viewed on the Open Science Framework repository for this project: https://osf.io/wq37x

### Procedure

All participants completed an online survey hosted at the JISC Online Surveys platform at baseline (‘T1’). First, participants provided basic demographic information, self-reporting whether they had a neurological, vestibular (including duration of dizziness symptoms) or psychiatric diagnosis, their education level, general health, number of medications, balance problems, number of falls in the past 12 months, and whether they require assistance for basic activities of daily living. As per previous related research,^13^ we also asked participants if they ever “experienced dizziness that was unrelated to alcohol or drug consumption or standing up too quickly”.

Participants then completed a battery of questionnaires. This included first the newly developed 11-item Balance-VQ. Participants were asked to score each item from 1 (never) to 5 (always), with respect to how they “typically feel in relation to their balance”. Next, they completed the Vertigo Symptom Scale (VSS)-short form.^14^ This 15-item scale assess the frequency of vertigo, dizziness and associated automatic arousal symptoms over the past month. Items are scored from 0 (never) to 4 (very often [most days]), with total scores thus ranging from 0 to 60. It has two subscales: One assessing vertigo symptoms (8-items) and another assessing autonomic arousal (7-items). Participants also completed the 7-item anxiety subscale of the Hospital Anxiety and Depression Scale (HADS-A) which assesses recent symptoms of anxiety.^15^ Finally, they completed the 7-item short version of the Falls Efficacy Scale-International^16^ to provide information on the degree of any concerns about falling experienced.

The first 125 participants were then invited to complete the Balance-VQ again 2 weeks later (‘T2’), for test-retest reliability. Here, they also reported whether they had experienced any falls, or serious worsening of balance or dizziness in the 2 weeks since first completing the Balance-VQ. This served as our anchor: only participants that had not fallen, and had not experienced serious worsening of balance/dizziness were included in the re-test analyses.

### Statistical Analysis

All data were analysed with SPSS and AMOS (version 28; IBM, Chicago, IL). Unless stated otherwise, alpha was set at *p*=.05. Figure 1 summarises the flow of the study and analyses, which broadly involved the following steps:

**Figure 1.**
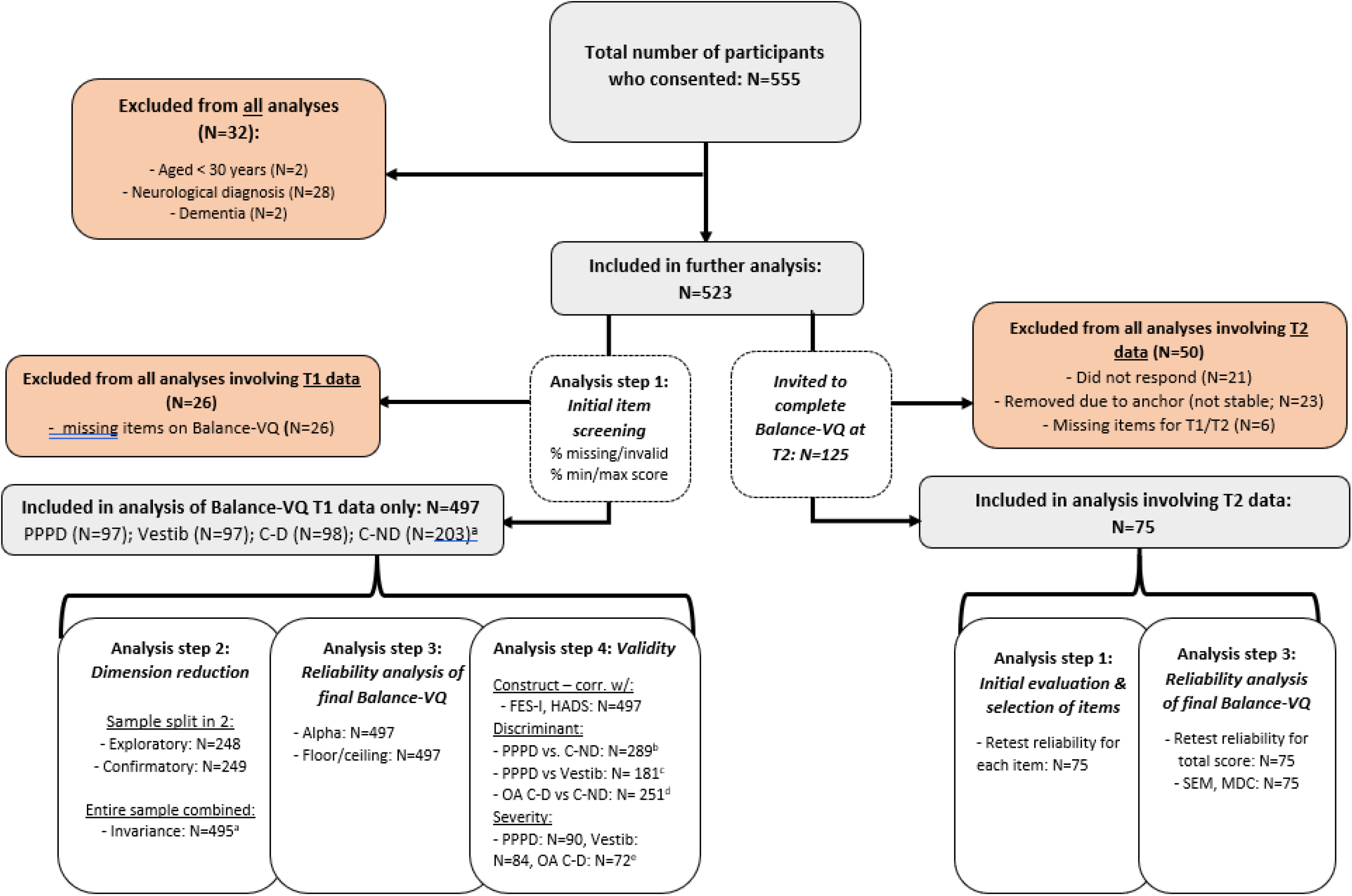
Study flow. **NB:** ^a^Two participants had failed to respond to the questions about self-reported dizziness. These have been removed only from analysis where subgrouping based on this variable was relevant; ^b-e^ For these regression analyses, some participants were excluded due to missing data for at least one of the control variables: ^b^ 5 PPPD group members and 6 C-ND group members excluded; ^c^ 5 PPPD and 8 vestibular group members excluded; ^d^ Only the control group participants aged ≥ 60 years were included, of which 5 older control-dizzy and 7 older control-not dizzy participants were excluded. ^e^ 7 PPPD, 13 vestibular, and 15 older adult C-D group members excluded; **Abbreviations:** C-D = Controls without diagnosed neuro-otological dysfunction, but with self-reported dizziness; C-ND: Controls without diagnosed neuro-otological dysfunction, and with no self-reported dizziness; OA = Older adults; PPPD = Persistent postural perceptual dizziness; Vestib = Vestibular diagnosis group

#### Step 1 – Screening of individual items

Balance-VQ items were considered for removal if: (i) there were a large number of missing (or multiple) responses (>5%) at T1; (ii) >50% of responses at T1 were the minimum or maximum score, or (iii) Individual item’s test-retest reliability was low (2-way, random effect, consistency single measures ICC<.5).^17^

#### Step 2 - Dimension reduction and validation

Next, exploratory factor analysis and subsequent confirmatory factor analysis were performed. Participants were first randomly allocated (using random.org, 50:50 ratio) to either an ‘exploratory’ or ‘confirmatory’ subsample. We then first performed exploratory analysis using principal axis factoring (varimax rotation) using the T1 Balance-VQ data for the ‘exploratory’ subgroup. Visual inspection of the scree plot was done to identify the number of latent factors. Items were considered for removal if they loaded on multiple factors, loaded insufficiently (<0.4)^18^ on a single factor, or if they exhibited low item-rest (r’s<0.3) and/or high inter-item correlations (r’s>.7). This was followed by confirmatory factor analysis using the T1 Balance VQ data of the ‘confirmatory’ subgroup (using maximum likelihood estimation).^19^ Items were constrained to load on the underlying factor(s) they had been associated with in the exploratory factor analysis. Pairs of error terms within a factor could co-vary if this improved model fit. We evaluated model fit using pre-defined criteria, ^20–22^ see Supplementary Material 3 for further details.

So-called “measurement invariance” was determined to assess whether the scale structure was similar across participant from the PPPD, vestibular and control groups with or without dizziness (i.e., 4 subgroups in total). For this analysis we used T1 Balance-VQ scores from the entire sample, and recommended criteria to evaluate changes in model fit for different levels of invariance.^23^ See Supplementary materials for details).

#### Step 3 - Reliability and measurement error

In this step, we evaluated the finalised Balance-VQ’s internal consistency (Cronbach’s alpha) and test-retest reliability (2- way, random effect, consistency, single measures ICC). For both alpha and ICC, we considered values >.70 to be satisfactory. Also, measurement error (SEM = SD + 2*√(1-ICC))^24^ and minimal detectable change were assessed on group and individual level (MDC_group_= SEM × 1.96 × √2/√n; MDC_individual_ = SEM × 1.96 × √2).^25^ Finally, floor and ceiling effects were determined (i.e., >15% of participants scoring lowest/highest possible summed scores^26^).

#### Step 4 - Construct validity

Construct validity was assessed by correlating (Spearman’s rho) Balance-VQ total scores with scores on (i) measures of balance-specific (Falls-Efficacy Scale) and (ii) generalised anxiety (HADS-anxiety). Construct validity would be evidenced if we would find significant weak to moderate correlations (.3-.5).

Further, we used logistic regressions to assess if Balance-VQ scores were predictive of group status for groups in which balance vigilance has been implicated as potential risk factor, when controlling for known confounding variables (age, gender, generalised anxiety symptoms, depression diagnosis, no of medications). Specifically, we assessed if people with higher Balance-VQ scores would have greater odds of (i) PPPD group membership versus the control-no dizziness group membership, (ii) PPPD group membership versus vestibular group membership, and (iii) experiencing dizziness vs no dizziness in daily life among control group members above 60 years of age (as discussed in the introduction, this is a group which frequently experiences ‘unexplained dizziness’^7, 8^). These analyses were done in two steps, i.e., only using Balance-VQ scores as independent variable for the first step, followed by the addition of control variables in a second step.

Finally, we conducted linear regressions to assess if Balance-VQ score was associated with dizziness severity (VSS total scores), when correcting for the same confounding variables as reported above. We did this analysis separately for the PPPD, vestibular, and control group (for the latter focusing on older adults (>60 years) with dizziness only).

#### Step 5 – ROC analysis to identify cut-off values

ROC analyses were conducted to determine optimal cut-off points to determine: (1) PPPD vs. controls (non-dizzy); (2) Older adult controls dizzy vs. older adult controls non-dizzy, and; (3) individuals meeting the cut-off point for anxiety using the HADS (≥11/21) vs. below this cut-off value, given the previously reported associations between anxiety and *general* bodily vigilance in balance disorders.^5^

#### Sample size considerations

We aimed for an overall sample of 500 participants, to allow for two samples of 200 participants each for the exploratory and confirmatory factor analyses, respectively (exceeding recommended subject-to-variable ratio of 10:1^27^). For test–retest reliability analysis, power analysis showed that a sample size of 60 “stable” participants would be more than sufficient to be able to detect an ICC of 0.80 with a 95% CI of .70-.90.

#### Missing data

Participants with any missing data for the Balance-VQ were excluded from all analyses (*N* = 26; See Supplementary Materials for information regarding missingness for individual Balance-VQ items). Missing data rates for Short FES-I, VSS-sf and HADS-A were 0.2%, 0.8% and 1.9%, respectively. As per recommendations,^16, 28^ missing items for the Short FES-I and the HADS-A were handled using the individual-mean imputation approach.^c^ As there are no guidelines for handling missing data for the VSS-sf, participants with any missing data were excluded from all analyses involving this scale (*N* = 33).

### Data availability and pre-registration

Analyses and data-handling procedures were pre-registered (https://osf.io/d9hxn?view_only=true and https://osf.io/abxec). Data relevant to this project can be found here: https://osf.io/x4zph/. This project page also contains a document that details (justification for) any major deviations from the registered analysis protocol: https://osf.io/tgh6c

## Results

### Participant Characteristics

Figure 1 summarises the flow of the study. In total, 555 participants completed the study at T1. Of these, 32 were excluded as they did not meet inclusion criteria.

Further, as stated earlier, 26 participants had missing data for 1 or more items of the Balance-VQ, and hence were excluded from all further analyses. Therefore, in total we included T1 data from 497 participants. Of these, 97 participants had a diagnosis of PPPD, and 97 had a current vestibular diagnosis other than PPPD^d^. While the remaining 303 control participants were without diagnosed neuro-otological dysfunction, 98 of these did self-report dizziness complaints in daily life (control-dizzy), with 203 control participants reporting no dizziness (control-not dizzy). Participant characteristics are summarised for each of these four participant groups in Table 1.

Seventy-five participants also completed the Balance-VQ at T2. Of these, 13 had PPPD, 13 had a vestibular diagnosis that was not PPPD, 14 were classified as control-dizzy, and 35 were classified as control-not dizzy. Overall, the retest sample’s characteristics were broadly similar to that of the overall (N=497) sample (See Supplemental Material 1 for a detailed overview).

### Initial screening and selection of items

We evaluated the performance of the individual items of the Balance-VQ. There were no clear issues with missing items (N=30 in total, N≤7 (1.3%) for separate items). Evaluation of scoring distribution and of reliability indices revealed potential issues with item 9 (minimum value for 58% of participants) and with item 11 borderline floor (42%) and poor re-test reliability (ICC=.549). These two items were therefore excluded from further analyses. Supplementary Material 2 summarises item-level analysis results.

### Dimension Reduction and Validation

Exploratory factor analysis on the 9 remaining items (items 1-8, and item 10), revealed a one-factor solution (explained variance=61.7%). With the exception of item 10, all items loaded on this factor (loadings≥.611; see Table 2). Item 10 was therefore removed from further analysis. Further, we removed items 1 and 8 due to very high inter-item correlations between item 1 and item 3 (*r*=.815), and between item 8 and items 1,3,5 (*r*’s=.751-.766). The analysis was run a second time without items 1, 8, and 10. Only one component was identified, explaining 66.9% of variance. All six items loaded highly on this component (.618-.850; see Table 2).

**Table 2.** Factor loadings for each item, presented separately for each of the two runs of the factor analysis.

| Item | RUN 1<sup>a</sup> | RUN 2<sup>b</sup><br>(after excluding<br>items 1, 3 and 10) |
| --- | --- | --- |
|  | Factor Loading<br>(explained variance 61.7%) | Factor Loading<br>(explained variance 66.9%) |
|  | *Factor 1* | *Factor 1* |
| 1. I closely monitor how steady my balance feels | **.827** | n/a |
| 2. I become alarmed by sudden or temporary changes in steadiness | **.722** | **.850** |
| 3. I am vigilant to small changes in how steady my balance feels | **.866** | **.830** |
| 4. I immediately know when my balance worsens | **.668** | **.618** |
| 5. When something happens that affects my balance, I am anxious to check how much my steadiness has decreased | **.783** | **.835** |
| 6. I worry about fluctuations in steadiness | **.751** | **.846** |
| 7. I avoid situations that I fear will affect my balance and make me less steady | **.611** | **.668** |
| 8. I keep careful track of how steady my balance feels | **.846** | n/a |
| 10. I remain calm in situations which worsen my balance | .071 | n/a |

<sup>a</sup>Kaiser-Meyer-Olkin assessment (KMO) = .917; all individual KMOs ≥ .749 (>0.5 threshold; Field 2018).
<sup>b</sup>KMO = .896; individual KMOs ≥ .882;

### Confirmatory Factor Analysis and Measurement Invariance

Overall, confirmatory factor analysis supported the one-factor structure identified in the exploratory factor analysis. The model demonstrated sufficient measurement invariance (see Supplemental Material 3), suggesting that the structure of the Balance-VQ is similar for the different populations tested: i.e., controls with or without dizziness experiences, people with a vestibular diagnosis, and people with a diagnosis of PPPD. Figure 2 presents the final Balance-VQ.

**Figure 2.**
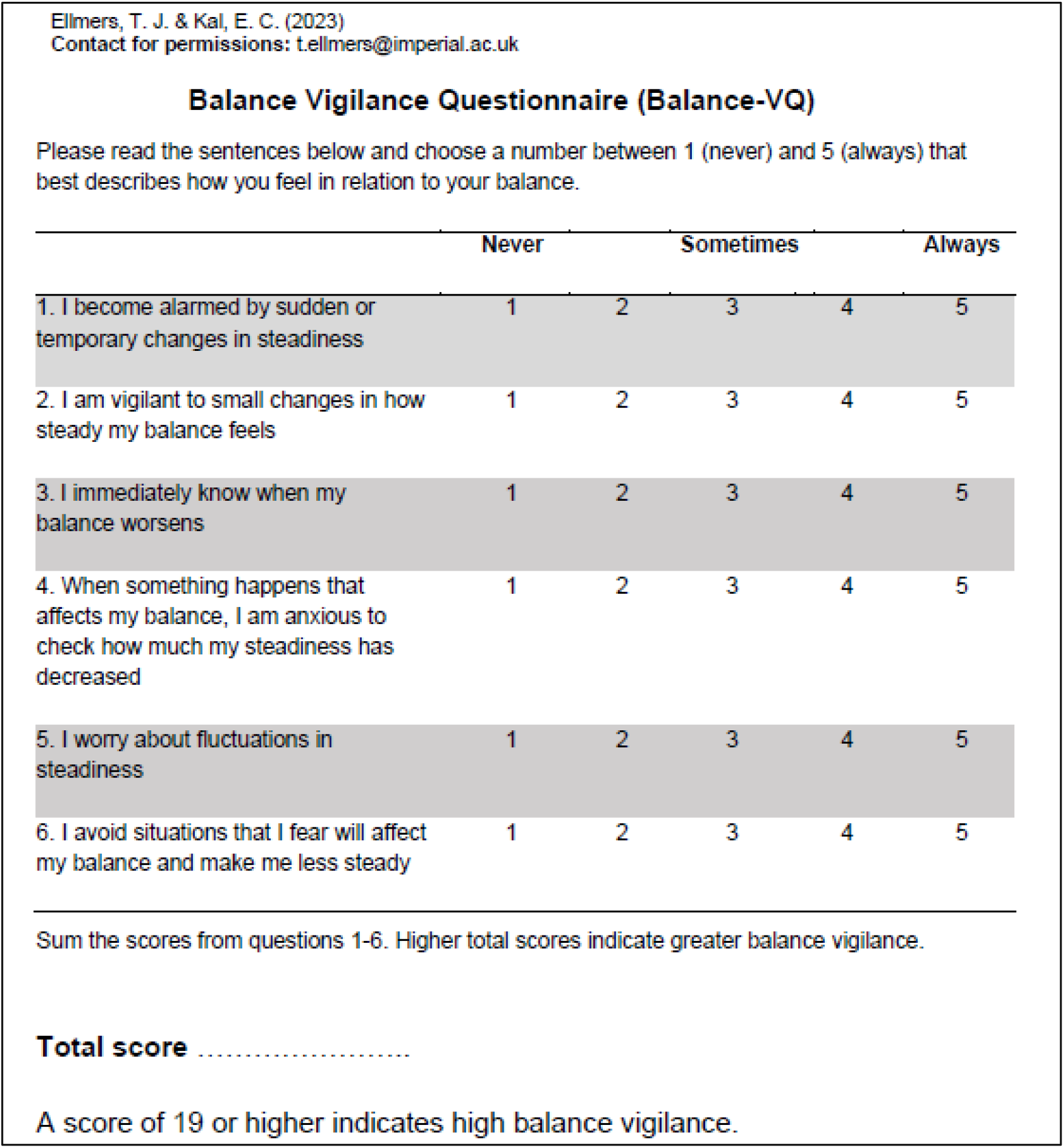
Final 6-item Balance Vigilance Questionnaire (Balance-VQ). Overall balance vigilance is scored by summing the scores for each items. Total scores range between 6-30, with higher scores indicating greater balance vigilance.

### Reliability and Measurement Error Analysis

The Balance-VQ showed excellent internal consistency (alpha=.905) and test-retest reliability (ICC=.797, 95%CI=[.697, .867]). The standard error of measurement was 2.93 points. Minimal detectable change values were 0.9 (group level) and 8.1 (individual level). There were no clear floor and ceiling effects: 7.6% (N=38) of individuals scored the minimal possible score (6 points), whereas 4.0% (N=20) scored the maximal possible score (30 points).

### Construct Validity Analysis

#### Correlations of Balance-VQ with other related constructs

Summed Balance-VQ scores correlated with Falls-Efficacy Scale scores (*r* = .624, *p* < .001, 95% CI = [.567, .674], N = 497) and generalised anxiety (HADS-Anxiety scores; *r* = .483, *p* < .001, 95% CI = [.412, .548], N = 497). Broadly speaking, this confirmed the hypothesis that the Balance-VQ measures related but distinct constructs compared to these outcome measures, although the correlation with FES scores was somewhat higher than anticipated.

#### Associations with Balance-VQ and participant group

Mean scores for the control-not dizzy, control-dizzy, PPPD, and vestibular groups at T1 are presented in Figure 3a. As we also made specific comparisons with the controls ≥ 60 years of age, their data can be observed in Figure 3b.

**Figure 3.**
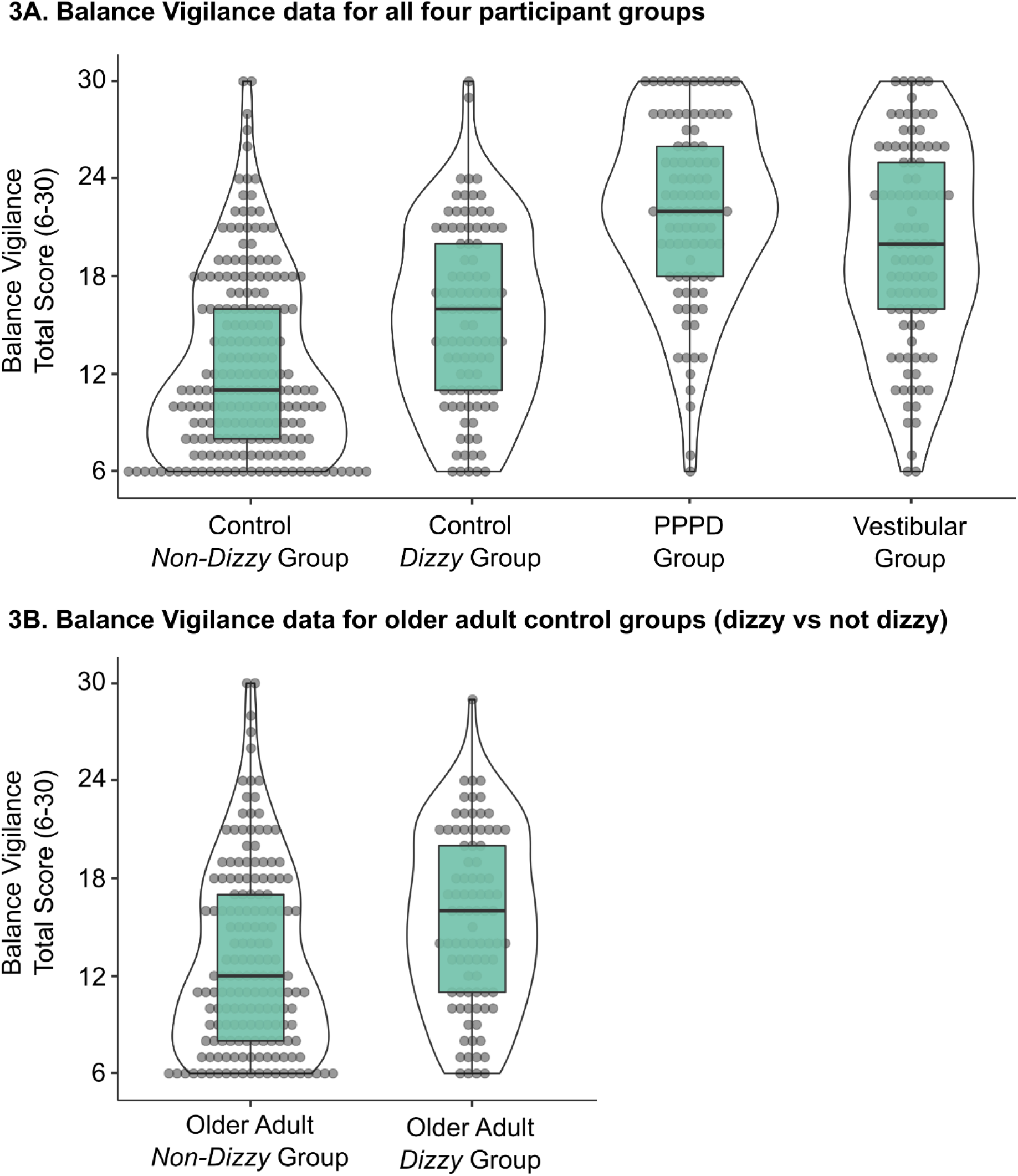
Total summed Balance Vigilance Scores (6-30) for all four participant groups (3A), as well as presented separately for the older adults (≥60 years of age; 3B) in the non-dizzy vs dizzy control groups. Note, the ‘Vestibular’ group consists of individuals diagnosed with a vestibular disorder *other* than PPPD, whilst the ‘Control Dizzy’ group consists of individuals without diagnosed neuro-otological dysfunction who nonetheless experience *some* degree of dizziness. Median, interquartile range and individual scores are presented.

For detailed results of the logistic regression analyses, please see Supplemental Materials 4. In short, higher Balance-VQ scores were associated with greater odds of being in the PPPD group vs the Control-not dizzy group even when controlling for potential confounding factors (OR: 1.34 95%CI=[1.23, 1.46]). Also, for older adult control participants, higher Balance-VQ scores were associated with greater odds of reporting dizziness in daily life (OR: 1.06, 95%CI=[1.01, 1.12]). However, although Balance-VQ scores were associated with greater odds of having PPPD rather than a Vestibular diagnosis (OR: 1.07 95%CI=[1.02, 1.13]), these scores no longer discriminated between the two groups when controlling for confounding factors (OR: 1.03, 95%CI=[0.97, 1.10]).

#### Associations with Balance-VQ scores and dizziness severity

Finally, in PPPD, Vestibular and older adult Control-dizzy groups, Balance-VQ scores were significantly associated with VSS scores – even when controlling for confounding variables (PPPD: β=0.83, 95%CI=[0.43, 1.22], p<.001; Vestibular: β=0.54, 95%CI=[0.20, 0.88], p=.002; Control-dizzy: β=0.33, 95%CI=[0.14, 0.52], p<.001). This association appeared the strongest for the PPPD group; not only with respect to the β value (adjusted for confounding variables) reported above, but also since adding the confounding variables did not significantly increase the model fit. See Table 3 for detailed results for each group.

**Table 3.**
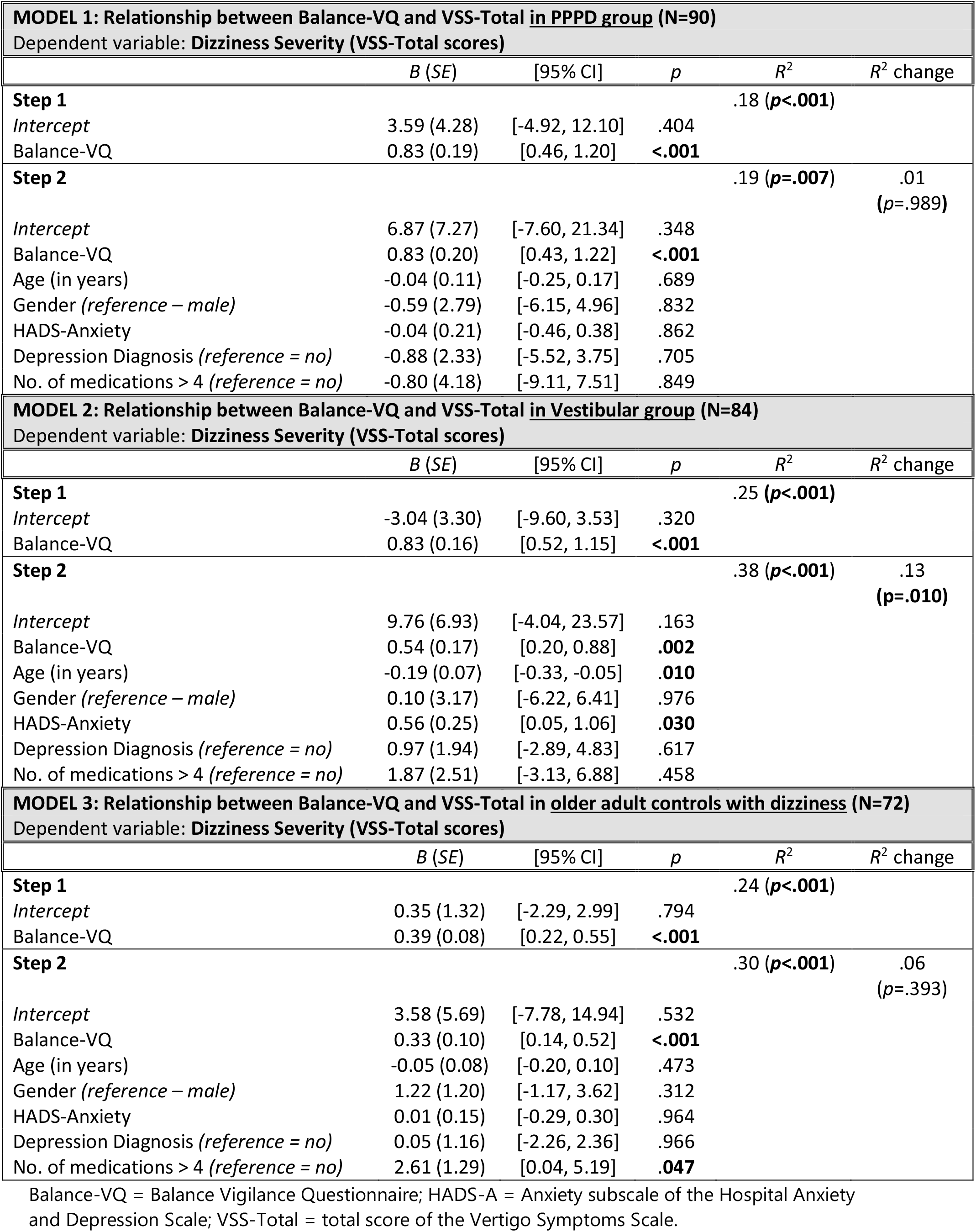
Linear regressions exploring the relationship between Balance-VQ and VSS-Total scores across the three groups (PPPD, ‘other’ vestibular disorder and older adult controls who reported experiencing dizziness in daily life).

| **MODEL 1: Relationship between Balance-VQ and VSS-Total in PPPD group (N=90)** |  |  |  |  |  |
| --- | --- | --- | --- | --- | --- |
| Dependent variable: Dizziness Severity (VSS-Total scores) |  |  |  |  |  |
|  | *B (SE)* | [95% CI] | *p* | *R*<sup>2</sup> | *R*<sup>2</sup> change |
| **Step 1** |  |  |  | .18 (*p*<.001) |  |
| *Intercept* | 3.59 (4.28) | [-4.92, 12.10] | .404 |  |  |
| Balance-VQ | 0.83 (0.19) | [0.46, 1.20] | <.001 |  |  |
| **Step 2** |  |  |  | .19 (*p*=.007) | .01<br>(*p*=.989) |
| *Intercept* | 6.87 (7.27) | [-7.60, 21.34] | .348 |  |  |
| Balance-VQ | 0.83 (0.20) | [0.43, 1.22] | <.001 |  |  |
| Age (in years) | -0.04 (0.11) | [-0.25, 0.17] | .689 |  |  |
| Gender (*reference – male*) | -0.59 (2.79) | [-6.15, 4.96] | .832 |  |  |
| HADS-Anxiety | -0.04 (0.21) | [-0.46, 0.38] | .862 |  |  |
| Depression Diagnosis (*reference = no*) | -0.88 (2.33) | [-5.52, 3.75] | .705 |  |  |
| No. of medications > 4 (*reference = no*) | -0.80 (4.18) | [-9.11, 7.51] | .849 |  |  |
| **MODEL 2: Relationship between Balance-VQ and VSS-Total in Vestibular group (N=84)** |  |  |  |  |  |
| Dependent variable: Dizziness Severity (VSS-Total scores) |  |  |  |  |  |
|  | *B (SE)* | [95% CI] | *p* | *R*<sup>2</sup> | *R*<sup>2</sup> change |
| **Step 1** |  |  |  | .25 (*p*<.001) |  |
| *Intercept* | -3.04 (3.30) | [-9.60, 3.53] | .320 |  |  |
| Balance-VQ | 0.83 (0.16) | [0.52, 1.15] | <.001 |  |  |
| **Step 2** |  |  |  | .38 (*p*<.001) | .13<br>(*p*=.010) |
| *Intercept* | 9.76 (6.93) | [-4.04, 23.57] | .163 |  |  |
| Balance-VQ | 0.54 (0.17) | [0.20, 0.88] | .002 |  |  |
| Age (in years) | -0.19 (0.07) | [-0.33, -0.05] | .010 |  |  |
| Gender (*reference – male*) | 0.10 (3.17) | [-6.22, 6.41] | .976 |  |  |
| HADS-Anxiety | 0.56 (0.25) | [0.05, 1.06] | .030 |  |  |
| Depression Diagnosis (*reference = no*) | 0.97 (1.94) | [-2.89, 4.83] | .617 |  |  |
| No. of medications > 4 (*reference = no*) | 1.87 (2.51) | [-3.13, 6.88] | .458 |  |  |
| **MODEL 3: Relationship between Balance-VQ and VSS-Total in older adult controls with dizziness (N=72)** |  |  |  |  |  |
| Dependent variable: Dizziness Severity (VSS-Total scores) |  |  |  |  |  |
|  | *B (SE)* | [95% CI] | *p* | *R*<sup>2</sup> | *R*<sup>2</sup> change |
| **Step 1** |  |  |  | .24 (*p*<.001) |  |
| *Intercept* | 0.35 (1.32) | [-2.29, 2.99] | .794 |  |  |
| Balance-VQ | 0.39 (0.08) | [0.22, 0.55] | <.001 |  |  |
| **Step 2** |  |  |  | .30 (*p*<.001) | .06<br>(*p*=.393) |
| *Intercept* | 3.58 (5.69) | [-7.78, 14.94] | .532 |  |  |
| Balance-VQ | 0.33 (0.10) | [0.14, 0.52] | <.001 |  |  |
| Age (in years) | -0.05 (0.08) | [-0.20, 0.10] | .473 |  |  |
| Gender (*reference – male*) | 1.22 (1.20) | [-1.17, 3.62] | .312 |  |  |
| HADS-Anxiety | 0.01 (0.15) | [-0.29, 0.30] | .964 |  |  |
| Depression Diagnosis (*reference = no*) | 0.05 (1.16) | [-2.26, 2.36] | .966 |  |  |
| No. of medications > 4 (*reference = no*) | 2.61 (1.29) | [0.04, 5.19] | .047 |  |  |

Balance-VQ = Balance Vigilance Questionnaire; HADS-A = Anxiety subscale of the Hospital Anxiety and Depression Scale; VSS-Total = total score of the Vertigo Symptoms Scale.

#### Cut-off values to identify high balance vigilance

Area under the curve scores were 0.88 for PPPD vs. controls (non-dizzy); (2) 0.63 for older adult controls dizzy vs. older adult controls non-dizzy, and; 0.78 for individuals meeting the cut-off point for anxiety using the HADS vs. below this cut-off value. Based on these analyses, we defined the cut-off point of ≥19/30 on the Balance-VQ to identify someone as having high balance vigilance.

## Discussion

The aim of this research was to develop (through patient input) a validated balance-specific measure of vigilance that comprehensively assesses the varied ways in which this construct may manifest. Our findings confirm that the Balance-VQ is a valid and reliable self-report instrument for assessing vigilance towards balance. A recent review exploring factors predicting the development of PPPD and chronic dizziness recommended that clinicians screen for bodily vigilance to identify the patients most at risk of developing symptoms of chronic dizziness following an acute vestibular insult.^4^ We propose that the short, 6-item Balance-VQ (with the established cut-off point of ≥19/30) would be well-suited for such purpose.

As hypothesised, Balance-VQ scores were higher in individuals diagnosed with PPPD, compared to controls (without dizziness), and scores predicted group membership even when controlling for confounding variables such as age, gender and anxiety. These findings support previous work highlighting the role of *generalised* bodily vigilance in the development and maintenance of PPPD.^1–5^ Although the present study was cross-sectional rather than prospective in nature, prospective designs have identified generic (i.e., not specific to balance) vigilant monitoring of bodily signals as a strong risk factor for the development of PPPD.^5^ Given the generic nature of the tools used to explore this relationship, we propose that the more specific assessment of *balance* vigilance developed in the present work may be an even more sensitive measure for predicting the development of chronic dizziness. This could then help identify which patients may be most likely to benefit from rehabilitation which specifically targets attention during balance; a strategy which seems particularly effective for improving symptoms in PPPD (see^29^). Future work should look to explicitly test the sensitivity of the Balance-VQ for predicting the development of chronic dizziness (particularly following an acute neuro-otological insult).

Whilst the specific mechanism through which balance vigilance may contribute to PPPD remains unknown, there is a wealth of evidence that describes how consciously attending sensory input can alter the way in which the brain processes these signals (e.g.,^30–32)^. Some researchers have hypothesised that heightened monitoring of balance in PPPD may therefore amplify the neural processing of discrepancies between anticipated and actual postural feedback signals (i.e., ‘prediction errors’).^2, 3^ This could then lead to individuals becoming aware of minor changes in postural sway which are always occurring (see^33^), but that typically take place outside of conscious awareness given that the ‘error signals’ are so low.^34^ As van den Bergh et al.^35^ write, “if there are no cues directing attention to the body, minor prediction errors may go unnoticed” (p. 195). Supporting this stance, recent research highlights that conditions which increase the amount of attention directed towards consciously monitoring balance lead to the cortical processing of minor changes in postural stability (changes which would otherwise be largely ignored at the cortical level).^36^

Similar mechanisms have also been proposed to contribute to the distorted perceptions of instability that are common in older adults^6, 37^ – particularly in those with ‘unexplained dizziness’.^7, 38^ In line with these suggestions, Balance-VQ scores were higher in older adults without diagnosed neuro-otological dysfunction but who nonetheless experience dizziness and perceive themselves to be unstable. A general sensitivity for changes in bodily signals may serve an adaptive purpose. But persistent *active* scanning in order to detect cues for one’s physical condition may change the way that the brain processes this information, serving to exacerbate symptom perception – particularly when this leads to pervasive worries when symptoms are detected.^39^ However, future research should look to scrutinise the specific mechanisms through which balance vigilance contributes to perceptions of instability and chronic dizziness across different populations.

Balance-VQ scores correlated to dizziness severity (as measured by the VSS-sf) in all three ‘dizzy’ groups: PPPD, diagnosed vestibular disorder other than PPPD, and the ‘Dizzy Control’ group (those without diagnosed neuro-otological dysfunction who nonetheless experience *some* degree of dizziness). However, based on confidence intervals and mean estimates, this association was strongest for the PPPD group. This could be meaningful, as it indicates that the relationship between dizziness and balance vigilance may differ across types of dizziness. Alternatively, it could be an artefact of the PPPD group having on average more severe dizziness. Future work should therefore look to explore this association further. Interestingly, Balance-VQ scores were also high in a number of control individuals without dizziness, with around 15% scoring above the identified cut-off point for high balance vigilance (≥19/30). Like other symptoms and factors associated with PPPD (e.g., visual motion sensitivity),^13^ this suggests that balance vigilance might exist on a spectrum in the general population. Whether or not high scores on the Balance-VQ then predisposes an individual to developing chronic dizziness following an acute neuro-otological insult should be explored in future work.

Although Balance-VQ scores were higher in the PPPD group compared to individuals with diagnosed vestibular disorders other than PPPD, Balance-VQ scores did not discriminate between the two groups when confounding factors were controlled for. However, a major limitation of the present work was the remote nature of the data collection (which took place due to COVID-19 restrictions with face-to-face testing). This meant that we were not able to conduct objective neuro-otological testing and rule out undiagnosed PPPD in the non-PPPD vestibular group. Future work should therefore look to compare Balance-VQ scores between individuals diagnosed with PPPD and a group of individuals diagnosed with a uniform peripheral vestibular disorder (e.g., bilateral vestibular hypofunction). Further, we recruited 4 quite distinct groups of participants, creating a relatively heterogeneous overall sample used for the factor analyses. However, our results showed that the scale demonstrates “measurement invariance”, meaning that the structure of the Balance-VQ holds similar across all groups (and that scores can be validly compared between these).

In summary, the short, 6-item Balance-VQ presented herein was shown to be valid and highly reliable tool to assess vigilance towards balance. Given the links between balance vigilance and chronic dizziness,^2, 4^ the Balance-VQ could be a useful to help identify patients most likely to benefit from rehabilitation that specifically targets attention during balance (see^29^). Future work should investigate the prospective utility of the scale for identifying those most at risk for developing chronic dizziness (e.g., PPPD).

## Acknowledgements

TJ Ellmers is supported by a Wellcome Trust Sir Henry Wellcome Postdoctoral Fellowship 222747/Z/21/Z.

The authors wish to thank: the patients and clinician PPI members who provided the crucial initial feedback on the scale; all the participants for volunteering their time to participate in the research; the social support groups and networks who agreed to distribute the survey amongst their members, and; the PPI members who read and provided feedback on earlier drafts on the manuscript.

## Supplementary Material 1. Characteristics of Test-Retest Sample (total N=75)

|  | Vestibular<br>(N = 13) | PPPD<br>(N = 13) | Controls<br>(no neuro-otological dysfunction; N = 49) |  |
| --- | --- | --- | --- | --- |
|  |  |  | Control-dizzy (N=14) | Control-not dizzy (N=35) |
| **Background / General Health** |  |  |  |  |
| Age in years (mean ± SD [range]) | 64.1 ± 11.2<br>[44-80] | 45.4 ± 11.3<br>[31-60]<sup>b</sup> | 70.5 ± 8.6<br>[59-85]<sup>a</sup> | 67.3 ± 15.4<br>[30-87] |
| Female gender (n; %) | 12 (92%) | 10 (77%) | 8 (57%) | 17 (49%) |
| Education Level – college / sixth form or above (n, %) | 12 (92%) | 10 (77%) | 11 (84%) | 30 (86%) |
| General health – self-reporting very good to excellent health (n; %) | 2 (15%) | 1 (8%) | 10 (71%) | 21 (60%) |
| Medication use >4 (n; %) | 2 (15%) | 2 (15%) | 1 (7%) | 5 (15%) |
| Peripheral neuropathy (n; %) | 0 (0%) | 0 (0%) | 0 (0%) | 1 (3%) |
| Diabetes (n; %) | 0 (0%) | 1 (8%) | 1 (7%) | 4 (11%) |
| **Physical functioning** |  |  |  |  |
| Falls in past 12 months (n; %) | 6 (46%) | 3 (23%) | 5 (36%) | 10 (29%) |
| Balance problems (n; %) | 11 (85%) | 10 (77%) | 6 (43%) | 5 (14%) |
| Walking aid (n; %) | 1 (8%) | 1 (8%) | 2 (14%) | 1 (3%) |
| ADL-assistance (n; %) | 1 (8%) | 1 (8%) | 0 (0%) | 0 (0%)<sup>a</sup> |
| **Psychological functioning** |  |  |  |  |
| Falls Efficacy Scale (7-28; mean ± SD [range]) | 13.2 ± 3.5<br>[8-20] | 13.0 ± 4.4<br>[8-21] | 9.6 ± 2.5<br>[7-15] | 9.4 ± 4.0<br>[7-27] |
| HADS-Anx (0-21; mean ± SD [range]) | 7.1 ± 3.8<br>[1-15] | 10.1 ± 5.5<br>[0-20] | 4.9 ± 3.1<br>[0-12] | 4.6 ± 3.8<br>[0-18] |
| Depression Diagnosis (n; %) | 4 (31%) | 7 (54%) | 3 (21%) | 3 (9%) |
| **Dizziness characteristics** |  |  |  |  |
| VSS – Total score (0-56; mean ± SD [range]) | 11.3 ± 8.0<sup>a</sup><br>[2-25] | 18.3 ± 10.1<br>[6-44] | 7.3 ± 4.6<sup>a</sup><br>[1-18] | 2.0 ± 2.2<sup>b</sup><br>[0-8] |
| VSS – Vertigo Subscale (0-28) (Mean ± SD [range]) | 6.8 ± 5.3<sup>a</sup><br>[1-18] | 11.5 ± 8.8<br>[1-30] | 2.9 ± 2.1<sup>a</sup><br>[0-7] | 0.5 ± 0.8<sup>b</sup><br>[0-2] |
| VSS – Arousal Subscale (0-28) (Mean ± SD [range]) | 4.5 ± 3.5<sup>a</sup><br>[0-12] | 6.9 ± 4.2<br>[2-16] | 4.4 ± 3.5<sup>a</sup><br>[1-12] | 1.6 ± 1.8<sup>b</sup><br>[0-7] |

**NB:** <sup>a</sup>1 missing value; <sup>b</sup>2 missing values;
**Abbreviations:** ADL: Activities of Daily Living; HADS-Anx: Anxiety subscale of the Hospital Anxiety Depression Scale; SD: Standard Deviation; VSS: Vertigo Symptom Scale;

## Supplementary Material 2. Results of initial screening of items

Results for step 1 of the analysis: item-level screening based on missing values, floor/ceiling effects, and retest reliability. Items 9 and 11 were excluded from further analysis based on this analysis. Note that item 11 technically met all required thresholds, but the combination of relatively low ICC value and a high proportion of people scoring minimum score made us decide to remove this item.

**Table S2.**
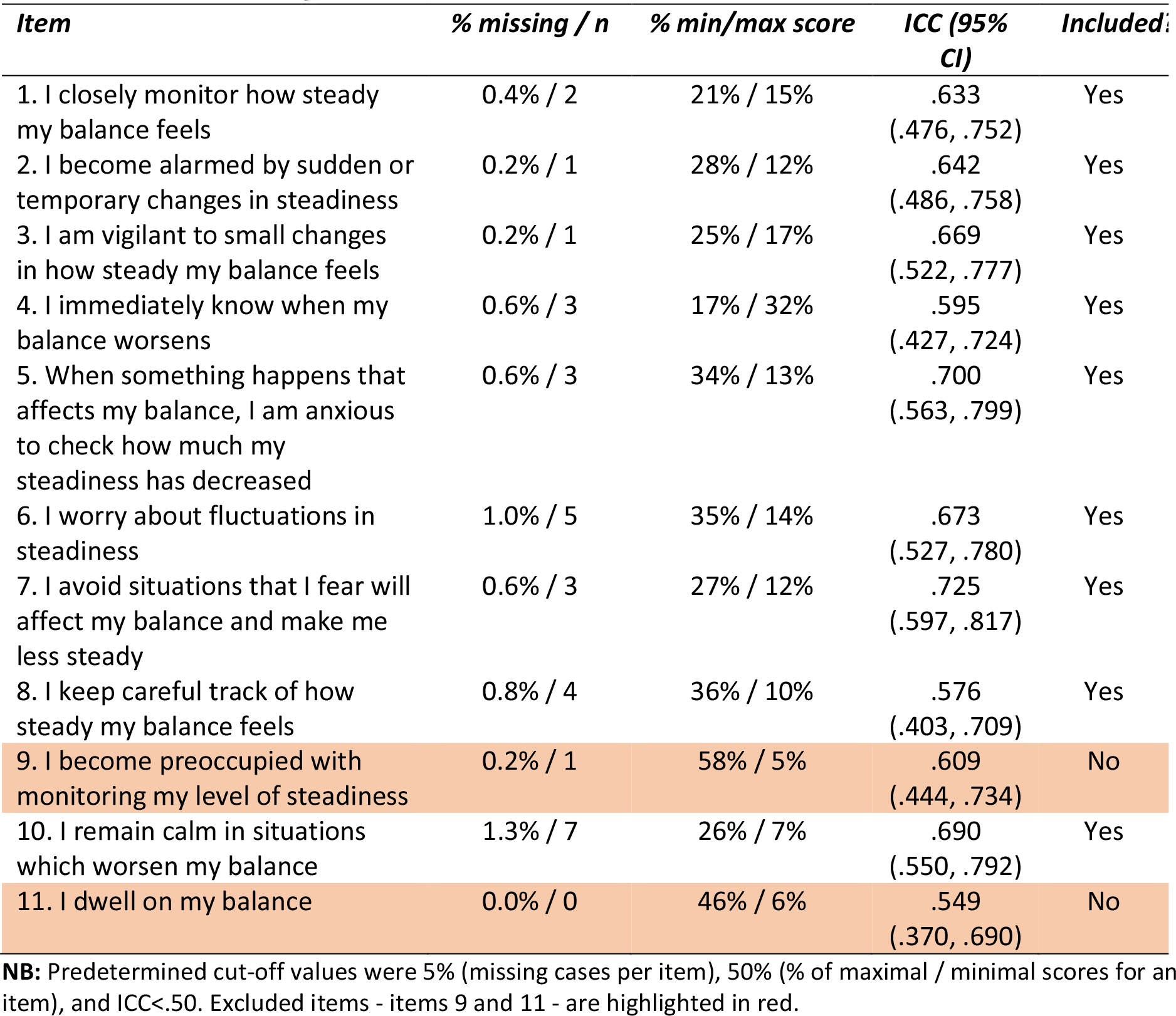
Initial screening of items.

| *Item* | *% missing / n* | *% min/max score* | *ICC (95% CI)* | *Included:* |
| --- | --- | --- | --- | --- |
| 1. I closely monitor how steady my balance feels | 0.4% / 2 | 21% / 15% | .633<br>(.476, .752) | Yes |
| 2. I become alarmed by sudden or temporary changes in steadiness | 0.2% / 1 | 28% / 12% | .642<br>(.486, .758) | Yes |
| 3. I am vigilant to small changes in how steady my balance feels | 0.2% / 1 | 25% / 17% | .669<br>(.522, .777) | Yes |
| 4. I immediately know when my balance worsens | 0.6% / 3 | 17% / 32% | .595<br>(.427, .724) | Yes |
| 5. When something happens that affects my balance, I am anxious to check how much my steadiness has decreased | 0.6% / 3 | 34% / 13% | .700<br>(.563, .799) | Yes |
| 6. I worry about fluctuations in steadiness | 1.0% / 5 | 35% / 14% | .673<br>(.527, .780) | Yes |
| 7. I avoid situations that I fear will affect my balance and make me less steady | 0.6% / 3 | 27% / 12% | .725<br>(.597, .817) | Yes |
| 8. I keep careful track of how steady my balance feels | 0.8% / 4 | 36% / 10% | .576<br>(.403, .709) | Yes |
| 9. I become preoccupied with monitoring my level of steadiness | 0.2% / 1 | 58% / 5% | .609<br>(.444, .734) | No |
| 10. I remain calm in situations which worsen my balance | 1.3% / 7 | 26% / 7% | .690<br>(.550, .792) | Yes |
| 11. I dwell on my balance | 0.0% / 0 | 46% / 6% | .549<br>(.370, .690) | No |

**NB:** Predetermined cut-off values were 5% (missing cases per item), 50% (% of maximal / minimal scores for an item), and ICC<.50. Excluded items - items 9 and 11 - are highlighted in red.

## Supplementary Material 3. Factor Analyses

Confirmatory factor analysis was performed using T1 data from the ‘confirmatory analysis subsample’ to evaluate model fit of a model where items 2-7 were constrained to load on one underlying factor/construct – i.e., the outcome from the exploratory analysis’ results. We evaluated model fit through assessment of: Standardised item-factor loadings, the chi-square statistic – both raw (χ^2^) and divided by its degrees of freedom (χ^2^ /df; both should be close to zero for good fit), goodness-of-fit and comparative fit indices (CFI; values>.95 and values>.90 indicate good and acceptable fit), standardized root mean squared residual (SRMR; values<.08 indicate good fit), and the root mean square error of approximation (RMSEA; values<.05 and values<.08 indicate good and acceptable fit, respectively).^1^

In an initial run, standardised item-factor loadings were all positive and high (.63-.91), with model fit indices showing mixed results (χ^2^(9)=52.869, p<.001; χ^2^/df=5.874; CFI=.957; GFI=.931; RMSEA=.140 [.105, .178]; SRMR=0.041). Inspection of the modification indices revealed that model fit could be substantially improved by allowing the residual error terms of items 3 and 4 to covary (MI=28.502). This was done in a second run. Item-factor loadings were again high and positive for each item (.60-.91), and model fit was largely satisfactory (χ^2^(8)=22.371, p=.004; χ^2^/df=2.796; CFI=.986; GFI=.970; RMSEA=.085 [.044, .128]; SRMR=0.028). Further inspection of the modification indices led us to the decision to also allow covariance between the residual error terms of items 4 and 5 (MI=6.620). This resulted in the final model as presented in Figure S3. Standardised item-factor loadings remained high (.59-.92), while model fit indices were now all acceptable to good (χ^2^(7)=15.558, p=.029; χ^2^/df=2.223; CFI=.992; GFI=.979; RMSEA=.070 [.021, .117]; SRMR=0.023).

**Figure S3.**
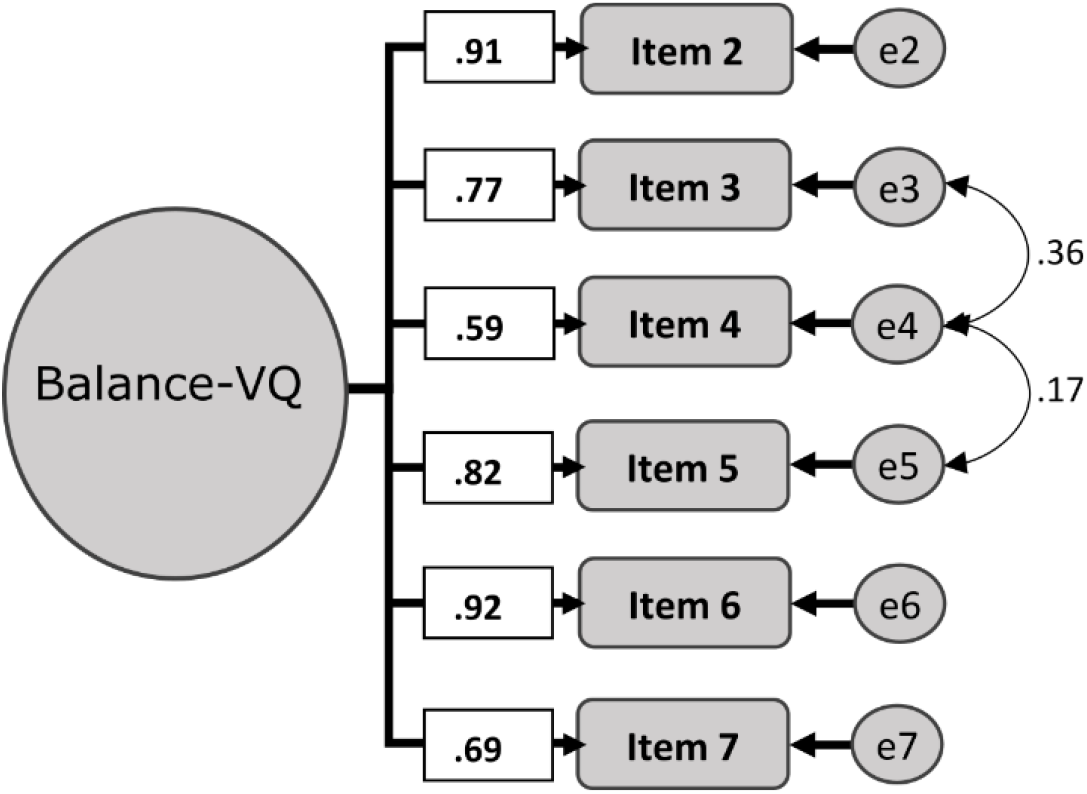
Final overall model yielded by the confirmatory factor analysis. Shown are the standardized item-factor loadings Abbreviated item numbers refer to the 6 selected items of the Balance-VQ). Also shown are the covariance between the residual error terms (abbreviated as ‘e’) of items 3 and 4, and 4 and 5.

## Measurement invariance testing

Table S3 shows the results of measurement invariance testing. To assess this, we evaluated model fit when item-factor loadings were free to differ between the 4 different participant subgroups (configural invariance), when item-factor loadings were equated across these groups (metric invariance), and when both the item-factor loadings and the intercepts of the model were equated across groups (scalar invariance). As shown in Table S3, we found evidence of sufficient configural, metric and scalar measurement invariance. Thus, the structure of the Balance-VQ seems similar across the different populations tested: i.e., controls with or without dizziness experiences, people with a vestibular diagnosis, and people with a diagnosis of PPPD.

**Table S3.**
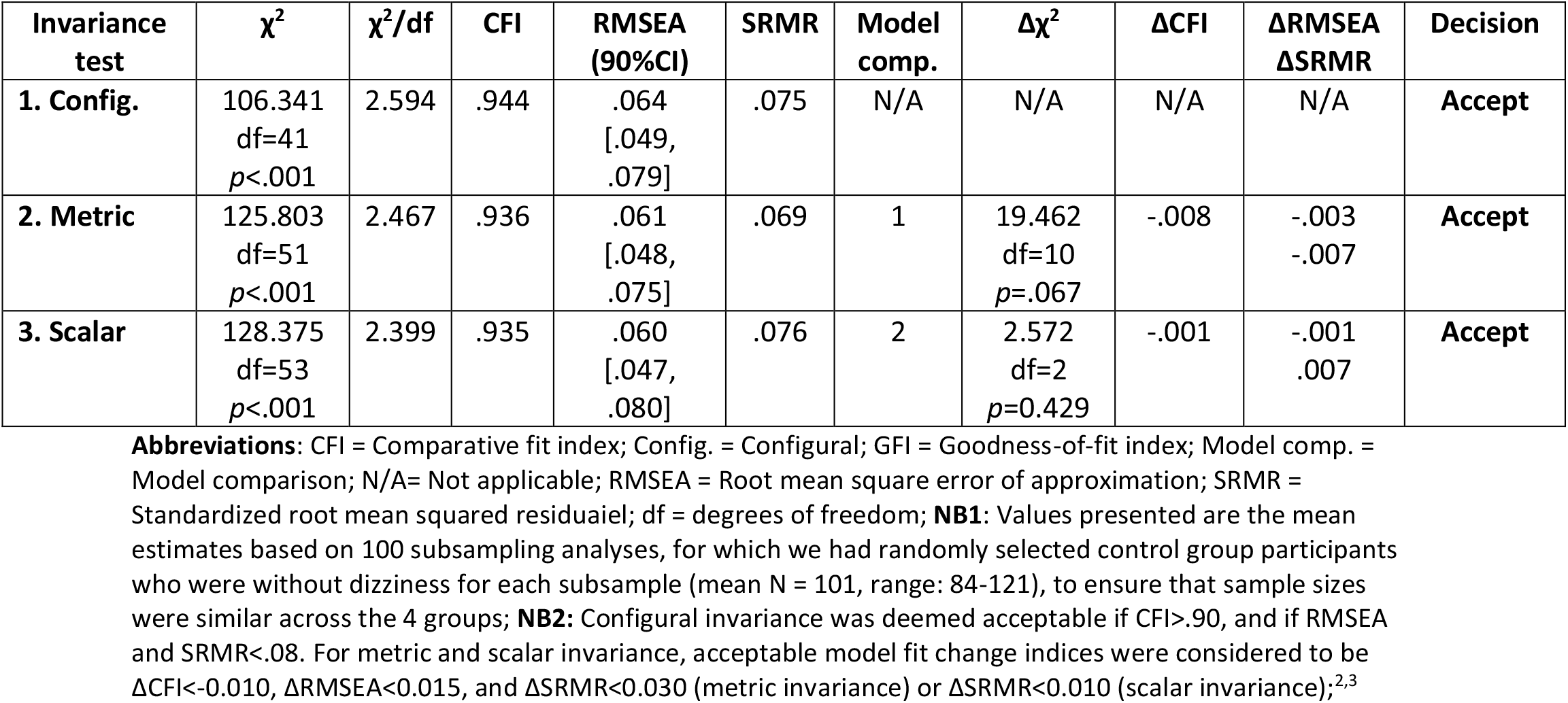
Results of measurement invariance testing.

| Invariance test | $\chi^2$ | $\chi^2/df$ | CFI | RMSEA (90%CI) | SRMR | Model comp. | $\Delta\chi^2$ | $\Delta CFI$ | $\Delta RMSEA$<br>$\Delta SRMR$ | Decision |
| --- | --- | --- | --- | --- | --- | --- | --- | --- | --- | --- |
| **1. Config.** | 106.341<br>df=41<br>$p<.001$ | 2.594 | .944 | .064<br>[.049,<br>.079] | .075 | N/A | N/A | N/A | N/A | **Accept** |
| **2. Metric** | 125.803<br>df=51<br>$p<.001$ | 2.467 | .936 | .061<br>[.048,<br>.075] | .069 | 1 | 19.462<br>df=10<br>$p=.067$ | -.008 | -.003<br>-.007 | **Accept** |
| **3. Scalar** | 128.375<br>df=53<br>$p<.001$ | 2.399 | .935 | .060<br>[.047,<br>.080] | .076 | 2 | 2.572<br>df=2<br>$p=0.429$ | -.001 | -.001<br>.007 | **Accept** |

**Abbreviations:** CFI = Comparative fit index; Config. = Configural; GFI = Goodness-of-fit index; Model comp. = Model comparison; N/A= Not applicable; RMSEA = Root mean square error of approximation; SRMR = Standardized root mean squared residual; df = degrees of freedom; **NB1:** Values presented are the mean estimates based on 100 subsampling analyses, for which we had randomly selected control group participants who were without dizziness for each subsample (mean N = 101, range: 84-121), to ensure that sample sizes were similar across the 4 groups; **NB2:** Configural invariance was deemed acceptable if CFI>.90, and if RMSEA and SRMR<.08. For metric and scalar invariance, acceptable model fit change indices were considered to be $\Delta CFI<0.010$ , $\Delta RMSEA<0.015$ , and $\Delta SRMR<0.030$ (metric invariance) or $\Delta SRMR<0.010$ (scalar invariance);<sup>2,3</sup>

## Supplemental Material 4. Results of the logistic and linear regression analysis

### Association between Balance-VQ scores and group status

Tables S4A-C present the outcomes for the logistic regression in which group status was predicted based on Balance-VQ score. In isolation – i.e., step 1 in each analysis – higher Balance-VQ scores were associated with greater odds of being classified in the PPPD group rather than in the Control-not dizzy group (OR: 1.32 95%CI=[1.24, 1.40]) or Vestibular group (OR: 1.07 95%CI=[1.02, 1.13]; Table S4A-B), as well as with greater odds of experiencing dizziness in daily life among older adults (OR: 1.08, 95%CI=[1.03, 1.13]; Table S4C).

When the control variables were added to each analysis – in step 2 – balance-VQ scores still differentiated between PPPD and Control-not dizzy groups (OR: 1.34 95%CI=[1.23, 1.46]), and between older adult Control-not dizzy and dizzy groups (OR: 1.06, 95%CI=[1.01, 1.12]; Tables S4A, S4C). However, higher balance VQ scores were no longer associated with significantly greater odds of belonging to the PPPD vs. vestibular group when control variables were added (OR: 1.03, 95%CI=[0.97, 1.10]; Table S4B).

### Association between Balance-VQ scores and dizziness severity

Tables S4D-F present the results of the association between Balance-VQ scores and VSS scores, separately for people with PPPD (Table S4D), people with a vestibular diagnosis (Table S4E), and older adult controls with dizziness in daily life (Table S4F). In all three analysis, Balance-VQ was associated with total VSS scores, and remained so when control variables were added to the regression models.

Please note that, for each group, a few participants could not be included due to missing values for one or more of the control variables and/or VSS score (PPPD: N=7, Vestibular: N=13, Control: N=15).

**Table S4A.** Results of logistic regression to determine association between Balance-VQ and PPPD status at T1 (PPPD = 1, N=92; vs Control-not dizzy = 0, N=197).^a^

|  | *p* | Odds Ratio [95% CI] |
| --- | --- | --- |
| **Step 1** |  |  |
| *Intercept* |  |  |
| Balance-VQ | **<.001** | 1.32 [1.24, 1.41] |
| **Step 2** |  |  |
| *Intercept* |  |  |
| Balance-VQ | **<.001** | 1.34 [1.23, 1.46] |
| Age (*in years*) | **<.001** | 0.90 [0.87, 0.94] |
| Gender (*reference = male*) | **.045** | 3.33 [1.03, 10.80] |
| HADS – Anxiety | **.022** | 1.13 [1.02, 1.26] |
| Depression diagnosis (*reference = no*) | **<.001** | 7.24 [2.26, 23.18] |
| No. of medications > 4 (*reference = no*) | 0.217 | 0.37 [0.08, 1.79] |

**NB:** <sup>a</sup> 5 PPPD and 6 control group members could not be included due to missing values for one or more of the control variables; **NB2:** Step 1: Nagelkerke $R^2=.53$ ; $\chi^2(1)=138.74$ , $p<0.001$ ; AUC = .89, Specificity = .85, Sensitivity = .75, cut-off: .375. Step 2: Nagelkerke $R^2=.79$ ; $\chi^2(6)=240.49$ , $p<0.001$ ; AUC = .97, Specificity = .90, Sensitivity = .89, cut-off: .30; **Abbreviations:** Balance-VQ = Balance Vigilance Questionnaire; CI = Confidence interval; HADS = Hospital Anxiety and Depression Scale; HC – no dizziness = Controls with no self-reported dizziness;

**Table S4B.** Results of logistic regression to determine association between Balance-VQ and PPPD status at T1 (PPPD = 1, N=92; vs Vestibular diagnosis = 0, N=89).^a^

|  | *p* | Odds Ratio [95% CI] |
| --- | --- | --- |
| **Step 1** |  |  |
| *Intercept* |  |  |
| Balance-VQ | **.007** | 1.07 [1.02, 1.13] |
| **Step 2** |  |  |
| *Intercept* |  |  |
| Balance-VQ | .362 | 1.03 [0.97, 1.10] |
| Age (*in years*) | **<.001** | 0.92 [0.89, 0.94] |
| Gender (*reference = male*) | .134 | 0.43 [0.14, 1.30] |
| HADS – Anxiety | .098 | 1.07 [0.99, 1.15] |
| Depression diagnosis (*reference = no*) | .084 | 0.52 [0.25, 1.09] |
| No. of medications > 4 (*reference = no*) | .457 | 0.66 [0.22, 1.99] |

**NB:** <sup>a</sup> 5 PPPD and 8 vestibular group members could not be included due to missing values for one or more of the control variables; **NB2:** Step 1: Nagelkerke $R^2=.06$ ; $\chi^2(1)=7.70$ , $p=0.006$ ; AUC = .61, Specificity = .58, Sensitivity = .50, cut-off: .53; Step 2: Nagelkerke $R^2=.41$ ; $\chi^2(6)=66.13$ , $p<0.001$ ; AUC = .83, Specificity = .74, Sensitivity = .73, cut-off: .475; **Abbreviations:** Balance-VQ = Balance Vigilance Questionnaire; CI = Confidence interval; HADS = Hospital Anxiety and Depression Scale;

**Table S4C.** Results of logistic regression to determine association between Balance-VQ and dizziness status at T1 (Control-dizzy (≥60 years) = 1, N=82; vs Control-not dizzy (≥60 years) = 0, N=160).^a^

|  | *p* | Odds Ratio [95% CI] |
| --- | --- | --- |
| **Step 1** |  |  |
| *Intercept* |  |  |
| Balance-VQ | **.002** | 1.08 [1.03, 1.13] |
| **Step 2** |  |  |
| *Intercept* |  |  |
| Balance-VQ | **.027** | 1.06 [1.01, 1.12] |
| Age (*in years*) | .221 | 0.97 [0.93, 1.02] |
| Gender (*reference = male*) | .270 | 1.48 [0.74, 2.99] |
| HADS – Anxiety | **.009** | 1.14 [1.03, 1.25] |
| Depression diagnosis (*reference = no*) | .601 | 1.23 [0.57, 2.65] |
| No. of medications > 4 (*reference = no*) | .835 | 0.92 [0.42, 2.00] |

**NB:** <sup>a</sup> 5 older control-dizzy and 7 older control-not dizzy group members could not be included due to missing values for one or more of the control variables; **NB2:** Step 1: Nagelkerke $R^2=.06$ ; $\chi^2(1)=9.85$ , $p=0.002$ ; AUC = .63, Specificity = .63, Sensitivity = .51, cut-off: .34; Step 2: Nagelkerke $R^2=.13$ ; $\chi^2(6)=23.45$ , $p<0.001$ ; AUC = .68, Specificity = .63, Sensitivity = .63, cut-off: .32; **Abbreviations:** Balance-VQ = Balance Vigilance Questionnaire; CI = Confidence interval; HADS = Hospital Anxiety and Depression Scale;

## Footnotes

a One-item (out of 18) *does* ask about the amount of attention directed towards feelings of dizziness. However, this scale is not validated for separation of individual items. Further, restricting assessment of balance vigilance to a single item hinders our ability to perform a fine-grained exploration of this construct.

b Although 98 out of 303 did report experiencing some degree of *“dizziness that is unrelated to alcohol or drug consumption, or standing up too quickly”*. We thus also separated control participants into a ‘dizzy’ and ‘non-dizzy’ group for certain analyses; see Statistical Analysis section for further information.

c Note, this approach is recommended in cases where participants are missing data for either a single-item (Short FES-I) or 3 items or fewer (HADS-A). These criterium were met for all missing data in the present research.

d Breakdown of self-reported diagnoses is as follows: Labyrinthitis/Vestibular neuritis: N=29; Meniere’s: N=19; Vestibular migraine: N=19; BPPV: N=13; Vestibular hypofunction: N=2; Mal de Debarquement: N=1; Multiple: 8; Unspecified vertigo: N=6

1 Hu L, Bentler PM. Cut-off criteria for fit indexes in covariance structure analysis: conventional criteria versus new alternatives. Struct Equ Modeling 1999; 6: 1–55. https://doi.org/10.1080/10705519909540118

2 Browne MW, Cudeck R. Alternative ways of assessing model fit. Sociol Method Res 1992; 21: 230–58. https://doi.org/10.1177%2F0049124192021002005

3 Chen FF. Sensitivity of goodness of fit indexes to lack of measurement invariance. Struct Equ Modeling 2007; 14: 464-504. https://doi.org/10.1080/10705510701301834

